# Characterizing cancer patterns in Okinawan vs. mainland Japanese Americans: The Multiethnic Cohort Study

**DOI:** 10.1101/2025.07.14.25331338

**Authors:** Samantha A Streicher, Cherie Guillermo, Song-Yi Park, Charleston WK Chiang, John Shepherd, Xin Sheng, David Bogumil, S Lani Park, Iona Cheng, Unhee Lim, Adrian A Franke, Daniel O Stram, David V Conti, Christopher A Haiman, Lynne R Wilkens, Loïc Le Marchand

**Affiliations:** Population Sciences in the Pacific Program, University of Hawaii Cancer Center, Honolulu, Hawai‘i, USA; Center for Genetic Epidemiology, Department of Population and Public Health Sciences, Keck School of Medicine, University of Southern California, Los Angeles, California, USA; Department of Epidemiology and Biostatistics, University of California, San Francisco, San Francisco, California, USA

**Keywords:** Okinawan, cancer, body composition, biomarkers

## Abstract

Differences in cancer rates have been documented in Japan between Okinawa and mainland Japan. Limited data exist on whether these differences are also present for established populations of Okinawans and mainland Japanese in the United States. Dimensionality reduction techniques for genetic data combined with Okinawan surnames were used to identify Multiethnic Cohort Japanese American participants (N=24,484) of Okinawan or mainland descent. Cox proportional hazards models were used to compare cancer incidence between Okinawan and mainland Japanese participants. Geometric means were examined on a subset of MEC participants for circulating blood biomarker levels (N=2,980) and body composition (N=399). The Okinawan cluster included 3,649 individuals and the mainland cluster included 19,611 individuals. Okinawan individuals were more likely to have a higher average body mass index and shorter stature, better diet quality score, higher total energy intake, more alcohol consumption among drinkers, and a history of never smoking compared to mainland Japanese (all p-values<0.0001). In multivariable adjusted models, Okinawan women were more likely to be diagnosed with breast cancer (HR=1.36, 95% CI=1.07-1.73) and Okinawan men were less likely to be diagnosed with aggressive prostate cancer (HR=0.67, 95% CI=0.51-0.87) compared to their mainland Japanese counterparts. In subsets of MEC participants, adiponectin levels were lower, and C-reactive protein levels, visceral adipose tissue area (VAT) and the VAT-to-subcutaneous adipose tissue area ratio were higher, in Okinawans compared to mainland Japanese (all p-values<0.05). Results in this US-based sample are consistent with recent trends of higher breast and lower prostate cancer incidence rates in Okinawans reported from Japan.

**Novelty and impact:** Cancer rate differences have been documented in Japan between Okinawa and mainland Japan; however, limited data exist on whether these differences are present in Japanese Americans of Okinawan or mainland descent. We report significant differences in breast cancer, prostate cancers, body composition, and obesity-related biomarkers for these two groups. Our findings suggest that these cancer risk disparities may not be solely due to lifestyle, but could be explained by body composition, genetics, or unmeasured factors.

## Introduction

Differences in cancer rates are documented in Japan between residents of Okinawa and the mainland Japanese prefectures^1^. For example, from 2016 through 2020, for the four most common cancer types, age-adjusted incidence rates for breast and colorectal cancers were higher in residents of Okinawa while those for prostate and lung cancers were lower in residents of Okinawa compared to mainland Japan^1^. Limited data exist on whether these differences are also present in Okinawans and mainland Japanese individuals who have migrated to the United States (US). Outside of Okinawa, Hawai‘i has one of the longest-established and largest Okinawan communities^2^. In 1885, the first group of mainland Japanese immigrants (mostly from southern Japan) arrived in Hawai‘i^2^. Fifteen years later (in 1900), the first small group of Okinawan immigrants arrived in Hawai‘i: From this time until the passage of the US Immigration Act in 1924, there were around 400 to 600 emigrants annually from Okinawa to Hawai‘i^2^: Today it is estimated that there are at least 45,000-50,000 Okinawans in Hawai‘i with smaller migrations to California^2,3^.

Okinawa, the southernmost of 47 prefectures in Japan, has a population of about 1.3 million people^4^. Until 1897, when Okinawa was annexed by Japan, Okinawa was an independent kingdom with unique foods, culture, language, music, and religion^4^. From 1945 until 1972, Okinawa was under US administration before being returned to Japan^5^. Although a few centenarian counts suggested exceptional longevity in Okinawan residents as early as 1950, it was not until 1975 that the Japan Ministry of Health included Okinawa in its life tables^5^. This inclusion revealed Okinawa’s leading position in all longevity indicators: life expectancy at birth, life expectancy at 65, and the proportion of nonagenarians and centenarians^5^. Because of their extraordinary life expectancy, the Okinawan prefecture was named one of five Longevity Blue Zones by famed National Geographic explorer Dan Buettner^6^. Unfortunately, the life expectancy advantage in the Okinawan population over the Japan’s national average began to decline after 1985, and by 2002 residents of Okinawa had lost their longevity edge, with life expectancy at birth falling closer to the Japan’s national average^7^. The post-World War II US military occupation of Okinawa is believed to have caused significant dietary and lifestyle changes causing sustained deleterious health effects for the Okinawan population^7,8^.

In 1991, researchers from Kuakini Medical Center in Honolulu, Hawai‘i reported no significant differences in risk of several cancer sites (i.e., mouth-esophagus, stomach, colon, rectum, lung, prostate, bladder, and lymphosarcoma/leukemia) between 1,109 Okinawan and 6,674 mainland Japanese men who immigrated or were children of immigrants to the US and had been followed since 1968 by the Honolulu Heart Study (HHS)^9^. While this work prompted the important research of examining distinct groups within the Japanese American population, the Honolulu Heart Study may have limited generalizability to Okinawans residing in the US today as it only included men, was relatively small and, since the men were born around 1900 to 1920, soon after the main Okinawan migration to Hawai‘i, these men may have continued to experience a more traditional Okinawan and less Westernized lifestyle.

In the current study, we examined differences in cancer incidence (breast, colorectal, prostate, and lung) among 24,484 Multiethnic Cohort (MEC) Japanese American participants of Okinawan or mainland Japanese descent, who were born from 1918 through 1948. We also assessed differences in obesity-related biomarkers and body composition phenotypes in subsets of Japanese American MEC participants of Okinawan or mainland Japanese descent^10^.

## Material and Methods

### Study design and participants

A prospective cohort analysis was conducted among participants enrolled in the MEC^11^, which was established in 1993-1996 to investigate the etiology of cancer and other chronic diseases^11^. It comprises over 215,000 participants recruited from Los Angeles County and Hawai‘i at the ages of 45-75^11^. Blood (or saliva on a subgroup) was collected as part of the MEC biorepository from participants starting in 1995, but mostly between 2001 and 2005. The five main self-reported ethnic groups represented in the MEC are African American, Japanese American, Latino, Native Hawaiian, and White^11^. In this study, we examined 24,484 participants from the MEC biorepository, who self-reported as Japanese American.

### Identification of Japanese Americans of Okinawan and mainland Japanese descent

The dimensionality reduction techniques, principal component analysis (PCA) followed by uniform manifold approximation and projection (UMAP), were applied to our sample using the 15,678 shared single nucleotide polymorphisms genotyped across 39 genotyping platforms, mostly Illumina platforms, among the 24,484 MEC Japanese Americans with blood or saliva samples (**Figures 1 and 2**). After the UMAP analysis, three large and three small genetic clusters emerged among the Japanese American population (**Figure 2**). The Okinawan, part- Okinawan and part-mainland Japanese, and mainland Japanese clusters were marked with men who had surnames frequently found in the Okinawan prefecture, and not in other Japan prefectures, according to a 2012 reference list generated by Shizuoka University^12^. To mark an Okinawan last name on the plots two criteria were used: 1) the surname was most frequent in the Okinawan prefecture by absolute number and 2) Over 80% of individuals with the surname were found to reside in the Okinawan prefecture compared to other Japanese prefectures^12^.

**Figure 1.**
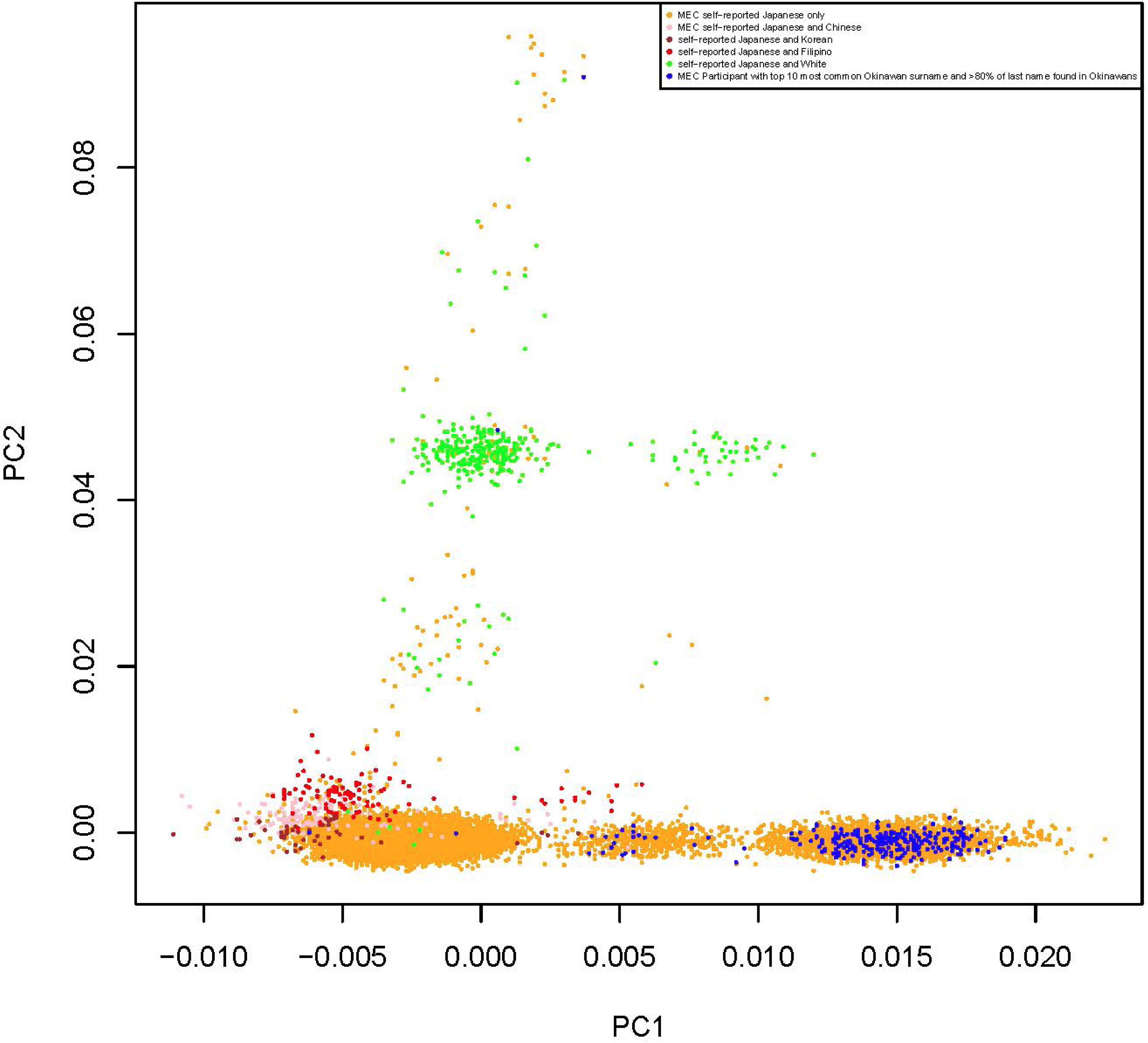
Principal component 2 vs. principal component 1 for 24,484 Japanese Americans.

**Figure 2.**
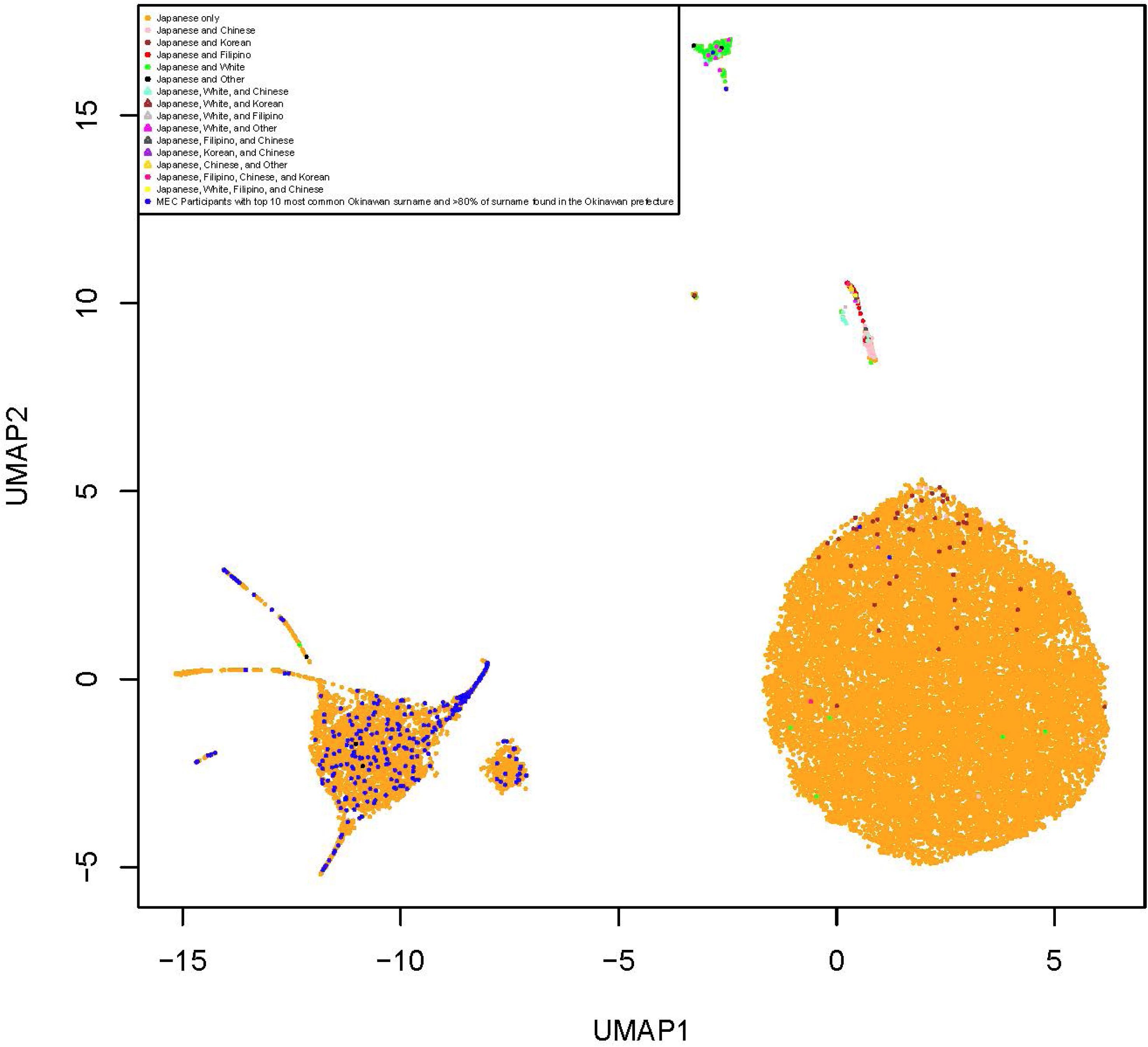
Uniform manifold approximation and projection (UMAP) 2 vs. UMAP 1 for 24,484 Japanese Americans with 6 principal components.

Historically, individuals from Okinawa were known to have unique surnames exclusive to this prefecture ^2^. Only male MEC participants were used as markers in order to avoid non-Okinawan married last names among women. On the UMAP plot, MEC participants in the large cluster with the vast majority of the labeled Okinawan surnames were identified as Japanese American individuals of Okinawan descent, participants in the large cluster void of labeled Okinawan surnames were identified as Japanese American individuals of mainland descent, and participants in the large cluster between the Okinawan and mainland Japanese cluster were identified as Japanese Americans of part-Okinawan and part-mainland descent (**Figure 2**). Other race and ethnicities were marked on the plots using a combination of every race and ethnicity self-reported on the MEC baseline questionnaire (**Figures 1 and 2**).

### Cancer outcomes

Incident invasive cancer diagnoses were identified via regular linkage to the Hawaii and California state Surveillance, Epidemiology, and End Results Program (SEER) cancer registries. Participants were followed after blood draw through December 31, 2019. Cases of breast, prostate, and lung cancers prevalent to blood draw (identified from the baseline questionnaire or by linkage to tumor registries) were excluded. Additionally, 2,274 women who self-reported premenopausal status from the baseline questionnaire were excluded from the breast cancer analysis. Aggressive prostate cancer was defined as regional or distant spread or localized with tumor grade three or higher.

### Obesity-related biomarkers

Obesity-related metabolic, hormonal, and inflammation dysfunction biomarkers were available from blood samples from a subset of 2,980 Japanese American individuals in the MEC biorepository. Blood samples were processed into components, and stored at -150°C. Plasma or serum concentrations were determined for total cholesterol (mg/dL), high-density lipoprotein (HDL) (mg/dL), low-density lipoprotein (LDL) (mg/dL), triglycerides (mg/dL), insulin (uIU/mL), glucose (mg/dL), leptin (pg/mL), adiponectin (µg/mL), sex-hormone binding globulin (SHBG) (nmol/L), total carotene (ng/mL), C-reactive protein (mg/L), and 25-hydroxyvitamin D3 (nmol/L) according to established methods.^13^ LDL was derived from the Friedewald equation using total and HDL cholesterol values and a valid range of triglyceride concentrations^14^.

### Body composition assessment

Body composition phenotype imaging for visceral adipose tissue (VAT) area, subcutaneous adipose tissue (SAT) area, percent liver fat, total fat mass, lean mass, muscle mass, and bone mass was available for 399 Japanese American MEC participants who also participated in the MEC-Adiposity Phenotype Study (APS)^10^. Extensive details regarding the design and imaging protocol, including quality control, have been previously published^10^. Briefly, in 2013-2016, MEC participants 60-77 years old who had not smoked in the past two years were examined after stratified enrollment on sex, race/ethnicity, and six body mass index (BMI) categories (18.5- 21.9, 22-24.9, 5-26.9, 27-29.9, 30-34.9, and 35-40 kg/m^2^)^10^. 3T magnetic resonance imaging (MRI) scanners (Siemens TIM Trio at the University of Hawaii and General Electric HDx at the University of Southern California) were used to assess abdominal fat distribution^10^. Whole-body composition was determined by dual-energy X-ray absorptiometry (DXA) (Hologic Discovery A fan-beam densitometer at the University of Hawaii and the University of Southern California)^10^.

### Statistical analysis

Participant characteristics were examined by mainland Japanese (n=19,611) or Okinawan (n=3,649) descent. The chi-square test was used to compare categorical variables and the Wilcoxon rank-sum test (Mann-Whitney U-test) to compare continuous variables. Cox proportional hazards analysis was used to calculate hazard ratios (HRs) and their 95% confidence intervals (95% CIs) with age as the time metric. Cancer outcomes for the most common cancers (breast, prostate, lung, colorectal) were modeled with observation start time at participants’ blood draw and end time at cancer diagnosis, death, or study end date of 12/31/2019, whichever came first. The primary independent variable of interest in all models was Okinawan versus mainland Japanese descent. Age-adjusted models were adjusted for age at blood draw as a strata variable. Multivariable models were further adjusted for known cancer risk factors assessed with the baseline MEC questionnaire administered from 1993 to 1996. For breast cancer, models included six additional covariates: BMI, age of menarche, age of menopause, hormone replacement use, number of children, and alcohol use. For colorectal cancer, models included 12 additional covariates: sex, BMI, pack-years, and aspirin use, multivitamin use, family history of colorectal cancer, hours in vigorous activity per day on average during the last year, daily intakes of red meat other than processed meat, processed red meat, vitamin D, dietary fiber, folate, and calcium. For prostate cancer, models were further adjusted for BMI and family history of prostate cancer in father or brother. In an exploratory analysis, prostate cancer models were additionally adjusted for the prostate-specific antigen (PSA) screening variable (ever vs. never) from the follow-up MEC questionnaire administered from 1998 to 2002 (N=9,768). For lung cancer, models included nine additional covariates: BMI, education, family history of lung cancer, physical activity, smoking status, average number of cigarettes per day, squared average number of cigarettes per day, number of years smoked, and number of years since quitting as time-dependent variables. The proportional hazards assumption was tested based on the interaction of time and exposure in a model; variables that did not meet the assumption were added as strata variables. To test for heterogeneity by sex, subsequent models with multiplicative variables between Okinawan/mainland Japanese descent and male/female were included in the regression. All statistical analyses were performed using SAS version 9.4 (Cary, North Carolina) and reported p-values are based on two-sided tests (summary alpha = 0.05). Analysis of covariance (ANCOVA) models were run with log transformed circulating biomarker values or body composition phenotype values and values were back transformed and reported as geometric means. Geometric means for circulating blood biomarker levels were reported after adjustment for 1) age of blood draw and sex and 2) age of blood draw, sex, and BMI. Geometric means were reported for body composition phenotypes after adjustment for age at entry into MEC-APS and sex, and VAT area was further adjusted for total fat mass. Values of p<0.05 were used to define statistical significance.

## Results

The Okinawan cluster included 3,649 individuals (15% of the MEC Japanese Americans) and the mainland Japanese cluster included 19,611 individuals (80% of the MEC Japanese Americans) from the UMAP plot (**Figures 1 and 2**). There were also smaller clusters of part-mainland Japanese and part-Okinawan individuals or part-mainland Japanese and part-white, Korean, Filipino, or Chinese (5% of the MEC Japanese Americans) who were not included in the analyses (**Figures 1 and 2**). Ninety-six percent of the Okinawan Americans and 94% of the mainland Japanese Americans were born in the US (**Table 1**). Okinawan individuals were more likely to have a higher BMI and shorter stature, a better diet quality score, a higher total energy intake, and a history of never smoking compared to mainland Japanese (all p-values<0.0001) (**Table 1**). Among alcohol drinkers, Okinawans consumed more alcohol compared to mainland Japanese (p-value<0.0001) (**Table 1**).

**Table 1.** Descriptive characteristics of Multiethnic Cohort (MEC) Japanese Americans source population, by mainland Japanese and Okinawan descent (N=23,994)

|  | mainland Japanese | Okinawan | p-value<sup>a</sup> |
| --- | --- | --- | --- |
| n (%) | 19,611 (80) | 3,649 (15) |  |
| Age at cohort entry (years), mean (SD) | 59.7 (8.7) | 59.8 (8.4) | 0.21 |
| Age at blood draw (years), mean (SD) | 69.0 (8.7) | 69.2 (8.5) | 0.17 |
| Sex, n (%) |  |  | 0.31 |
| Male | 9,388 (47.9) | 1714 (47.0) |  |
| Female | 10,223 (52.1) | 1935 (53.0) |  |
| Place of birth, n (%) |  |  | <0.0001 |
| United States | 18,402 (94.0) | 3,489 (95.8) |  |
| Japan (including Okinawa) | 1,097 (5.6) | 139 (3.8) |  |
| Other (including Europe, Central or South America, China, Korea, Philippines) | 83 (0.42) | 15 (0.41) |  |
| Place of recruitment, n (%) |  |  | <0.0001 |
| Hawai'i | 15,587 (79.5) | 3,290 (90.2) |  |
| Los Angeles | 4,024 (20.5) | 259 (9.8) |  |
| BMI (kg/m<sup>2</sup>), median (IQR) | 24.2 (19.6-28.8) | 25.0 (20.4-29.6) | <0.0001 |
| Weight (lbs), median (IQR) | 140 (120-160) | 140 (122-160) | 0.143 |
| Height (inches), median (IQR) | 63.7 (61-66) | 63 (60-65) | <0.0001 |
| Alcohol beverage intake status, n (%) |  |  |  |
| Drinker | 7,709 (40.4) | 1,439 (40.6) | 0.91 |
| Non-drinker | 11,352 (59.6) | 2,110 (59.4) |  |
| Alcohol beverage intake among drinkers (grams/day), median (IQR) | 101.8 (23.7-383.7) | 128.4 (24.9-511.6) | <0.0001 |
| Healthy Eating Index, median (IQR) | 66.9 (52.0-81.8) | 68.5 (53.3-82.1) | <0.0001 |
| Total energy intake (kcal/day) median (IQR) | 1,912.5 (1,480-2,480) | 1,976.1 (1,520 - 2,550) | <0.0001 |
| Smoking status, n (%) |  |  | <0.0001 |
| Never smoker | 9,958 (51.1) | 2,012 (55.6) |  |
| Former smoker | 7,624 (39.1) | 1,298 (35.9) |  |
| Current smoker | 1,912 (9.8) | 307 (8.5) |  |
| Pack-years among ever smokers, median (IQR) | 14.2 (6.2, 27.5) | 14.2 (6.2, 27.5) | 0.37 |
| Moderate to vigorous physical activity, median (IQR) | 0.82 (0.36-1.43) | 0.82 (0.36-1.54) | 0.42 |

<sup>a</sup>P-values were calculated using chi-square tests for categorical variables and Mann-Whitney U tests for continuous variables

Post-menopausal Okinawan women were significantly more likely to be diagnosed with breast cancer (age-adjusted HR=1.39, 95% CI=1.10-1.75) compared to postmenopausal mainland Japanese women (**Table 2**). This association remained even after adjusting for additional known breast cancer risk factors (i.e., BMI, age of menarche, age of menopause, hormone replacement use, number of children, and alcohol use) (HR=1.36, 95% CI=1.07-1.73) (**Table 2**). Okinawan men were significantly less likely to be diagnosed with aggressive prostate cancer (age-adjusted HR=0.67, 95% CI=0.51-0.87) (**Table 2**) compared to mainland Japanese ancestry individuals.

**Table 2.** Cox proportional hazards models for incident invasive breast, colorectal, prostate, and lung cancer in the MEC biocohort according to Okinawan vs. mainland Japanese origin^a^.

|  | Overall (N=22,267) |  |  |  |  | Women (n=11,763) |  |  |  |  | Men (n=10,504) |  |  |  |  |  |
| --- | --- | --- | --- | --- | --- | --- | --- | --- | --- | --- | --- | --- | --- | --- | --- | --- |
|  | Incidence, n | Total, n | Age-adjusted HR (95% CI)<sup>b</sup> | Age- and smoking-adjusted HR (95% CI) | Multivariable adjusted HR (95% CI) | Incidence, n | Total, n | Age-adjusted HR (95% CI)<sup>b</sup> | Age- and smoking-adjusted HR (95% CI) | Multivariable adjusted HR (95% CI) | Incidence, n | Total, n | Age-adjusted HR (95% CI)<sup>b</sup> | Age- and smoking-adjusted HR (95% CI) | Multivariable adjusted HR (95% CI) | P-heterogeneity by sex (male/female) for the multivariable adjusted model |
| **Breast Cancer<sup>c</sup>** |  |  |  |  |  |  |  |  |  |  |  |  |  |  |  |  |
| mainland Japanese | - | - | - | - | - | 334 | 7,351 | ref | - | ref | - | - | - | - | - | - |
| Okinawan |  |  |  |  |  | 89 | 1,421 | 1.39<br>(1.10-1.75) |  | 1.36 (1.07-1.73) |  |  |  |  |  |  |
| **Colorectal Cancer<sup>d</sup>** |  |  |  |  |  |  |  |  |  |  |  |  |  |  |  |  |
| mainland Japanese | 379 | 18,803 | ref | - | ref | 172 |  | ref | - | ref | 207 |  | ref | - | ref | 0.86 |
| Okinawan | 79 | 3,464 | 1.13<br>(0.88-1.45) |  | 1.12<br>(0.87-1.45) | 38 |  | 1.19<br>(0.84-1.69) |  | 1.19 (0.80-1.78) | 41 |  | 1.09<br>(0.8-1.52) |  | 1.08<br>(0.76-1.52) |  |
| **Prostate Cancer<sup>e</sup>** |  |  |  |  |  |  |  |  |  |  |  |  |  |  |  |  |
| mainland Japanese | - | - | - | - | - | - | - | - | - | - | 849 | 8,422 | ref | - | ref | - |
| Okinawan |  |  |  |  |  |  |  |  |  |  | 139 | 1,565 | 0.86<br>(0.72-1.03) |  | 0.87<br>(0.73-1.04) |  |
| **Aggressive Prostate Cancer<sup>e</sup>** |  |  |  |  |  |  |  |  |  |  |  |  |  |  |  |  |
| mainland Japanese | - | - | - | - | - | - | - | - | - | - | 462 | 8,422 | ref | - | ref | - |
| Okinawan |  |  |  |  |  |  |  |  |  |  | 59 | 1,565 | 0.67<br>(0.51-0.87) |  | 0.67<br>(0.51-0.87) |  |
| **Lung Cancer<sup>f,g</sup>** |  |  |  |  |  |  |  |  |  |  |  |  |  |  |  |  |
| mainland Japanese | 460 | 19,063 | ref | ref | ref | 177 | 9,996 | ref | ref | ref | 283 | 1,644 | ref | ref | ref | 0.055 |
| Okinawan | 65 | 3,509 | 0.75<br>(0.58-0.98) | 0.82<br>(0.63-1.06) | 0.81<br>(0.62-1.05) | 21 | 1,865 | 0.62<br>(0.40-0.98) | 0.66<br>(0.42-1.05) | 0.67 (0.43-1.07) | 44 | 9,067 | 0.83<br>(0.61-1.15) | 0.92<br>(0.67-1.28) | 0.90<br>(0.65-1.25) |  |

<sup>a</sup>Age was used as the time metric variable starting follow up at age of blood draw and ending follow up at age of cohort exit
<sup>b</sup>Age-adjusted models were adjusted for age at cohort entry (<50, 50–54, 55–59, 60–64, 65–69, ≥70 years) as a strata variable
<sup>c</sup>Multivariable-adjusted model for breast cancer was further adjusted for body mass index (<25, 25–29.9, ≥30 kg/m<sup>2</sup>) as a strata variable and age of menarche (<12, 13-14,>15), age of menopause (<45, 45-49, 50-54,>55, surgical menopause), hormone replacement use (yes, no), number of children (none,1,2-3,>=4), and alcohol use (drinks/day) as a covariates
<sup>d</sup>Multivariable-adjusted model for colorectal cancer was further adjusted for sex (male, female) body mass index (<25, 25–29.9, ≥30 kg/m<sup>2</sup>), and pack-years (number of cigarettes smoked per day x number of years smoked) as strata variables, and aspirin use (never,former,current), multivitamin use (yes, no), family history of colorectal cancer (yes,no), hours in vigorous activity per day (on average during the last year), red met other than processed meat (grams/day), processed red meat (grams/day), vitamin D use (IU/day), dietary fiber intake (grams/day), and dietary folate equivalents (micrograms) as covariates
<sup>e</sup>Multivariable-adjusted model for prostate cancer was further adjusted for body mass index (<25, 25–29.9, ≥30 kg/m<sup>2</sup>) as a strata variable and family history of prostate cancer (yes family history of prostate cancer in father or brother vs. no family history of prostate cancer in father or brother) as a covariate
<sup>f</sup>Age- and smoking-adjusted model for lung cancer was further adjusted for smoking status (never, former, current), average number of cigarettes per day, squared average number of cigarettes per day, number of years smoked (as a time-dependent variable), and number of years since quitting (as a time-dependent variable) as covariates
<sup>g</sup>Multivariable-adjusted model for lung cancer was further adjusted for body mass index (<25, 25–29.9, ≥30 kg/m<sup>2</sup>), education (<12th grade, vocational school, some college, >=college), family history of lung cancer (yes, no) as strata variables and physical activity (hours of moderate or vigorous activity per day), smoking status (never, former, current), average number of cigarettes per day, squared average number of cigarettes per day, number of years smoked (as a time-dependent variable), and number of years since quitting (as a time-dependent variable) as covariates

Similarly, this association did not change when adjusting for additional known prostate cancer risk factors (i.e., BMI and family history of prostate cancer in father or brother) (HR=0.67, 95% CI=0.51-0.87) or when further adjusting for PSA screening for early detection of prostate cancer (HR=0.68, 95% CI=0.52-0.89) (**Table 2 and Supplementary Table 1**). Okinawans were also less likely to be diagnosed with lung cancer (HR=0.75, 95% CI=0.58-0.98); however, this association was no longer significant when adjusting for smoking history (HR=0.82, 95% CI=0.63-1.06) (**Table 2**). There were no significant differences in risk of colorectal or all-stage prostate cancer between Okinawan and mainland Japanese individuals (**Table 2**). There were no significant interactions by sex for either colorectal or lung cancer (**Tables 2 and 3**).

**Table 3.**
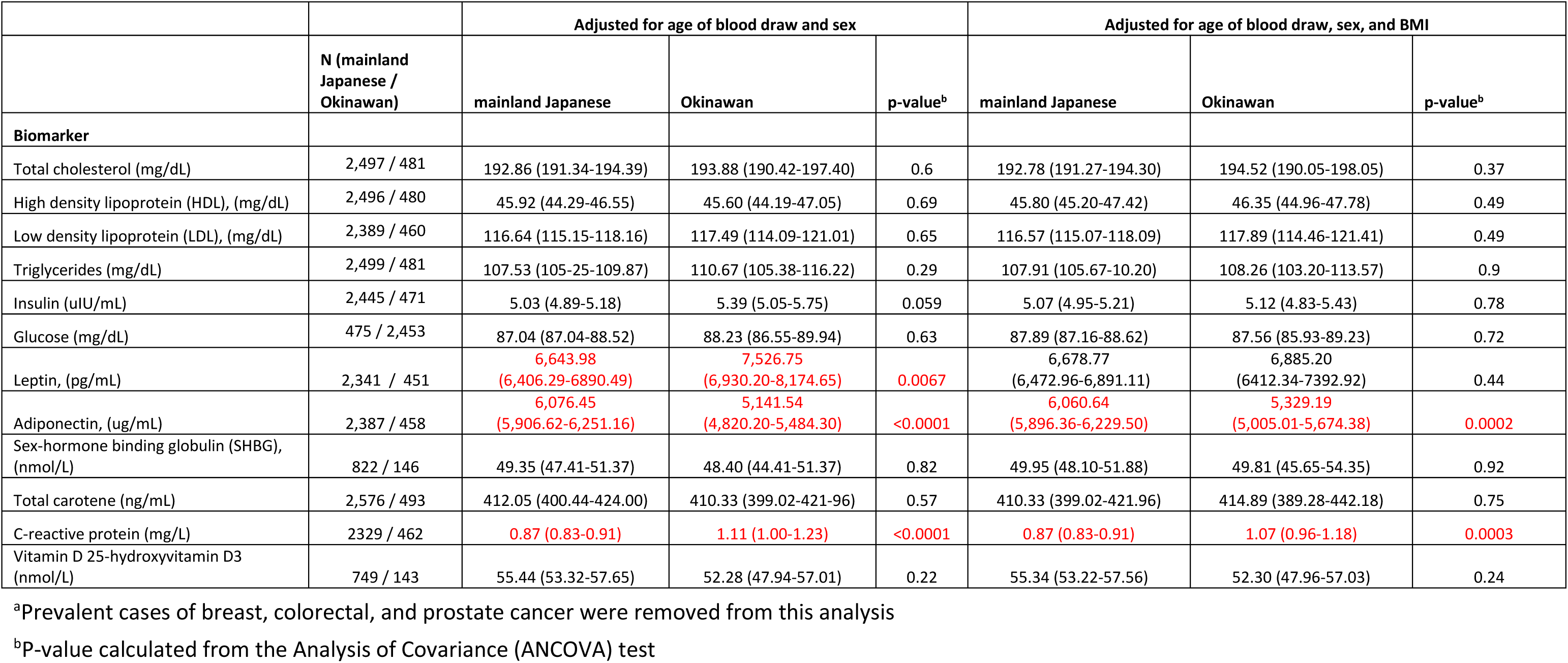
Geometric means of Obesity-related circulating biomarkers according to Okinawan vs. mainland Japanese descent^a^.

|  | N (mainland Japanese / Okinawan) | Adjusted for age of blood draw and sex |  |  | Adjusted for age of blood draw, sex, and BMI |  |  |
| --- | --- | --- | --- | --- | --- | --- | --- |
|  |  | mainland Japanese | Okinawan | p-value<sup>b</sup> | mainland Japanese | Okinawan | p-value<sup>b</sup> |
| **Biomarker** |  |  |  |  |  |  |  |
| Total cholesterol (mg/dL) | 2,497 / 481 | 192.86 (191.34-194.39) | 193.88 (190.42-197.40) | 0.6 | 192.78 (191.27-194.30) | 194.52 (190.05-198.05) | 0.37 |
| High density lipoprotein (HDL), (mg/dL) | 2,496 / 480 | 45.92 (44.29-46.55) | 45.60 (44.19-47.05) | 0.69 | 45.80 (45.20-47.42) | 46.35 (44.96-47.78) | 0.49 |
| Low density lipoprotein (LDL), (mg/dL) | 2,389 / 460 | 116.64 (115.15-118.16) | 117.49 (114.09-121.01) | 0.65 | 116.57 (115.07-118.09) | 117.89 (114.46-121.41) | 0.49 |
| Triglycerides (mg/dL) | 2,499 / 481 | 107.53 (105.25-109.87) | 110.67 (105.38-116.22) | 0.29 | 107.91 (105.67-10.20) | 108.26 (103.20-113.57) | 0.9 |
| Insulin (uIU/mL) | 2,445 / 471 | 5.03 (4.89-5.18) | 5.39 (5.05-5.75) | 0.059 | 5.07 (4.95-5.21) | 5.12 (4.83-5.43) | 0.78 |
| Glucose (mg/dL) | 475 / 2,453 | 87.04 (87.04-88.52) | 88.23 (86.55-89.94) | 0.63 | 87.89 (87.16-88.62) | 87.56 (85.93-89.23) | 0.72 |
| Leptin, (pg/mL) | 2,341 / 451 | 6,643.98<br>(6,406.29-6890.49) | 7,526.75<br>(6,930.20-8,174.65) | 0.0067 | 6,678.77<br>(6,472.96-6,891.11) | 6,885.20<br>(6412.34-7392.92) | 0.44 |
| Adiponectin, (ug/mL) | 2,387 / 458 | 6,076.45<br>(5,906.62-6,251.16) | 5,141.54<br>(4,820.20-5,484.30) | <0.0001 | 6,060.64<br>(5,896.36-6,229.50) | 5,329.19<br>(5,005.01-5,674.38) | 0.0002 |
| Sex-hormone binding globulin (SHBG), (nmol/L) | 822 / 146 | 49.35 (47.41-51.37) | 48.40 (44.41-51.37) | 0.82 | 49.95 (48.10-51.88) | 49.81 (45.65-54.35) | 0.92 |
| Total carotene (ng/mL) | 2,576 / 493 | 412.05 (400.44-424.00) | 410.33 (399.02-421.96) | 0.57 | 410.33 (399.02-421.96) | 414.89 (389.28-442.18) | 0.75 |
| C-reactive protein (mg/L) | 2329 / 462 | 0.87 (0.83-0.91) | 1.11 (1.00-1.23) | <0.0001 | 0.87 (0.83-0.91) | 1.07 (0.96-1.18) | 0.0003 |
| Vitamin D 25-hydroxyvitamin D3 (nmol/L) | 749 / 143 | 55.44 (53.32-57.65) | 52.28 (47.94-57.01) | 0.22 | 55.34 (53.22-57.56) | 52.30 (47.96-57.03) | 0.24 |

<sup>a</sup>Prevalent cases of breast, colorectal, and prostate cancer were removed from this analysis
<sup>b</sup>P-value calculated from the Analysis of Covariance (ANCOVA) test

Okinawan individuals with blood biomarkers assayed from the MEC biorepository had significantly higher levels of age- and sex-adjusted insulin (5.39 vs. 5.03 uIU/mL), leptin (7,527 vs. 6,644 pg/mL) and C-reactive protein (1.11 vs. 0.87 mg/L) and significantly lower levels of adiponectin (5,142 vs. 6,076 ug/mL) compared to mainland Japanese individuals (**Table 3**).

After further adjustment for BMI, levels of C-reactive protein remained significantly higher (1.07 vs. 0.87 mg/L) and levels of adiponectin remained significantly lower (7,125 vs. 7,731 ug/mL) (**Table 3**). Okinawan individuals who participated in the MEC-APS had a significantly shorter stature for both total height (62.0 vs. 63.5 inches) and sitting height (33.3 vs. 33.8 inches) compared to mainland Japanese, after adjustment for age and sex. MEC-APS Okinawan participants had a larger VAT area (154 vs 140 cm^2^), after adjustment for age, sex, and total fat mass, and a higher VAT-to-SAT ratio (1.02 vs. 0.88), after adjustment for age and sex, compared to mainland Japanese participants (**Table 4)**.

**Table 4.** Geometric means (95% confidence limits) of body composition phenotypes from the Multiethnic Cohort-Adiposity Phenotype Study (MEC-APS) according to Okinawan vs. mainland Japanese descent.

|  | **mainland Japanese**<br>**(n=327)** | **Okinawan (n=72)** | **p-value<sup>e</sup>** |
| --- | --- | --- | --- |
| **Body composition phenotype** |  |  |  |
| Body Mass Index (kg/m<sup>2</sup>)<sup>a</sup> | 25.71 (25.30-26.12) | 26.56 (25.67-27.47) | 0.088 |
| Body Mass Index, categorical |  |  |  |
| 18.5-21.9 kg/m<sup>2</sup> | 46 (13.9%) | 9 (12.5%) | 0.073 |
| 22.0-24.9 kg/m<sup>2</sup> | 92 (27.8%) | 19 (26.4%) |  |
| 25.0-26.9 kg/m<sup>2</sup> | 70 (21.2%) | 10 (13.9%) |  |
| 27.0-29.9 kg/m<sup>2</sup> | 76 (23.0%) | 15 (20.8%) |  |
| 30.0-34.9 kg/m<sup>2</sup> | 37 (11.2%) | 16 (22.2%) |  |
| 35.0-40.0 kg/m<sup>2</sup> | 6 (1.8%) | 3 (4.7%) |  |
| Weight (lbs)<sup>a</sup> | 146.97 (144.37-149.62) | 145.13 (139.74-150.73) | 0.55 |
| Height (inches)<sup>a</sup> | 63.47 (63.24-63.70) | 62.05 (61.57-62.53) | <0.0001 |
| Sitting height (inches)<sup>a</sup> | 33.79 (33.65-33.93) | 33.27 (32.99-33.57) | 0.0018 |
| Total fat mass (kg)<sup>a,b</sup> | 19.74 (19.16-20.35) | 19.53 (18.31-20.83) | 0.76 |
| Visceral adipose tissue area (cm<sup>2</sup>)<sup>a,c,d</sup> | 140.19 (135.13-145.44) | 153.94 (142.37-166.44) | 0.034 |
| Subcutaneous adipose tissue area (cm<sup>2</sup>)<sup>a,c</sup> | 168.36 (161.98-174.98) | 160.61 (147.96-174.35) | 0.31 |
| Visceral adipose tissue-to-subcutaneous adipose tissue ratio<sup>a,c</sup> | 0.88 (0.84-0.92) | 1.02 (0.93-1.12) | 0.005 |
| Percent Liver fat<sup>a,e</sup> | 5.03 (4.66-5.43) | 5.97 (5.07-7.02) | 0.063 |
| Lean mass (kg)<sup>a,b</sup> | 44.79 (44.13-45.46) | 44.00 (42.64-45.42) | 0.32 |
| Muscle mass (kg)<sup>a,b</sup> | 17.91 (17.62-18.20) | 17.42 (16.84-18.02) | 0.15 |
| Bone mass index (kg/height<sup>2</sup>)<sup>a,b</sup> | 0.79 (0.77-0.80) | 0.80 (0.77-0.80) | 0.4 |

<sup>a</sup>Adjusted for sex (male, female) and age at entry into the MEC-APS
<sup>b</sup>Total fat mass, lean mass, muscle mass, and bone mass were dual-exergy X-ray absorptiometry (DXA) measurements
<sup>c</sup>Visceral adipose tissue area, subcutaneous adipose tissue area, and percent liver fat were magnetic resonance imaging (MRI) measures
<sup>d</sup>Additionally adjusted for total fat mass (kg)
<sup>e</sup>P-value calculated from the Analysis of Covariance (ANCOVA) test

## Discussion

To our knowledge, this is the first study to examine within group heterogeneity in cancer risk for both men and women in a Japanese American population. Our findings reveal a significantly higher risk of postmenopausal breast cancer and a significantly lower risk of aggressive prostate cancer among Japanese Americans of Okinawan compared to mainland Japanese descent who were born between 1918 and 1948 (∼95% born in the US). While the Japan Ministry of Health calculated age-adjusted cancer incidence rates and we calculated multivariable-adjusted cancer hazard rates, both findings from the Japan Ministry of Health and our findings show higher rates for breast and colon cancer and lower rates for prostate and lung cancer when comparing residents of the Okinawan to mainland Japanese prefectures^1^.

Despite historically low cancer rates across all cancer sites reported for residents of Okinawa compared to mainland Japan, it is notable that recent patterns of incident cancer for the four most common cancer sites are comparable in Japan and in the US when comparing Okinawans to mainland Japanese^1^. Although a westernized diet and lifestyle was introduced at different times to the populations of the Okinawan prefecture (around 1950), mainland Japan (around 1970), and Okinawan and mainland Japanese immigrants to the US (early 1900s), it is possible that sufficient time has now elapsed in both Japan and the US to now observe higher rates for some cancer sites in both countries^7^. There is likely a combination of genetics, diet, and other lifestyle and environmental factors driving both the historically low rates of cancer and the modern higher rates of common cancers in these populations. For breast cancer, established risk factors (e.g., reproductive and hormonal risk factors, BMI, alcohol use) did not alter the higher risk of post- menopausal breast cancer in MEC Okinawans compared to mainland Japanese participants.

Recently in MEC, we reported that the frequency of a genetic variant significantly associated with the ratio of VAT to abdominal area was higher in Okinawan women compared to mainland Japanese women^15^. In the present study, we report that MEC Okinawan participants had a higher total fat mass-adjusted VAT area and VAT-to-SAT area ratio compared to mainland Japanese participants. VAT, the fat the surrounds internal abdominal organs, is an emerging independent risk factor for breast cancer with both genetic and environmental contributions^13,15,16^. It is possible that Okinawans compared to mainland Japanese, both in Japan and the US, are more predisposed than mainland Japanese and other populations to accumulate adiposity as VAT when exposed to a westernized diet and lifestyle^10^. VAT may be a stronger predictor of breast cancer risk than the genetic architecture of the disease or other unmeasured environmental factors, driving the recent higher risk of breast cancer observed in Okinawans compared to mainland Japanese. In comparison, there are few established risk factors for prostate cancer. For prostate cancer, it is plausible that the genetic architecture of the disease is a stronger predictor compared to environmental factors or the interplay of these factors.

In the MEC questionnaires, respondents were asked “What is your ethnic or racial background? (Mark all that apply)” with the response for Japanese American being “Japanese (includes Okinawan)”. Therefore, until now the MEC Japanese American population has been examined as one group. However, with recent genetic data for the 24,484 MEC Japanese Americans with blood samples, we were able to use dimensionality reduction techniques to expose the fine-scale structure in this population and examine within group heterogeneity for cancer risk.

Over 30 years ago, the Honolulu Heart Study was used to explore differences in cancer risk between self-reported Okinawan (n=1,109) and mainland Japanese (n=6,674) men in Hawaii^9^. These men were registered for military service between 1940 and 1942, interviewed and examined between 1965 and 1968, and followed up for cancer incidence with linkage to the Hawaii Tumor Registry and the Hawaii Department of Health for 19 years^9^. No statistically significant differences in site-specific cancer incidence between the Okinawan and mainland Japanese men were reported. However, age-adjusted incidence rates per 1,000 for stomach (24.4 vs. 33.5), rectal (11.2 vs 16.9), and prostate (32.8 vs. 43.3) cancers appeared at that time to be lower in Okinawans compared to mainland Japanese men^9^.

In our study, when comparing anthropometric measures and biomarker levels, we observed significantly higher levels for BMI, leptin, and C-reactive protein and lower levels of adiponectin for Okinawans compared to mainland Japanese Americans. These observed differences in biomarker levels are likely due to a greater adiposity in Okinawans, especially for visceral adipose tissue area, which is known to be more metabolically active compared to other fat areas^17^. While no other US study has compared underlying risk factors in disease incidence for Okinawans and mainland Japanese Americans, in Japan two large studies, the Japan Public Health Center-based Prospective Study (JPHC) and Biobank Japan (BBJ), have explored differences in anthropometric measures and biomarker levels between residents of Okinawa and mainland Japan^18,19^. The JPHC recruited over 100,000 participants throughout Japan, from Okinawa in the South to Ninohe in the North, with a first cohort recruitment for participants aged 40 to 59 years in 1990 and a second cohort recruitment for participants aged 40 to 69 years in 1993^20^. At baseline, five year follow-up, and ten year follow-up, residents of the Okinawan prefecture had a higher percentage of overweight (≥25 kg/m^2^) and obese (≥30 kg/m^2^) individuals compared to residents from mainland Japanese prefectures (e.g., Okinawan vs. mainland Japanese residents at baseline aged 50-54 percent overweight was 45.2% vs. 23.5% and percent obese was 4.8% vs. 1.5%)^20^.

BBJ recruited over 200,000 patients with at least one of 47 diseases (e.g., malignant tumors, respiratory diseases, and metabolic diseases) from 12 medical centers throughout Japan, including some in the Okinawa prefecture^21^. When comparing the same phenotypes that we examined in our study, BBJ observed higher blood circulating triglyceride levels, blood circulating C-reactive protein levels, BMI, and body weight and lower blood circulating HDL levels and height for Okinawans compared to mainland Japanese ^19^. It is possible that we did not observe differences in HDL, triglycerides, or body weight between mainland Japanese and Okinawans because Okinawan and mainland Japanese MEC participants were born, raised, and continue to live in a similar western environment, whereas Okinawan residents in Japan were introduced to a western diet and lifestyle earlier than mainland Japanese^2,22^.

In summary, we observed a significantly higher risk of postmenopausal breast cancer and a significantly lower risk of aggressive prostate cancer in MEC participants of Okinawa compared to mainland Japanese descent. These cancer risk patterns are similar to cancer incidence rates observed in residents from the prefecture of Okinawa compared to other prefectures in mainland Japan. This study suggests that these differences in cancer risk may not be due to lifestyle alone, but may be explained by biological differences related to body fat distribution, underlying genetics, or other unmeasured factors between the two populations. More extensive research is warranted to better understand the interplay between the traditional Okinawan diet, lifestyle factors, and genetics in this historically long-lived population^23^.

## Supporting information

prostate cancer risk with PSA screening

## Data availability statement

The data that supports the finding of this study are available upon request through the MEC online request system (https://www.uhcancercenter.org/for-researchers/mec-data-sharing).

## Ethics statement

All MEC participants provided written informed consent and the study was approved by the institutional review boards (IRBs) at the University of Hawaii (CHS-#17200), University of Southern California (#HS-12-00623), and University of California, San Francisco (#17–23399) in agreement with the 1975 Helsinki Declaration.

## Funding Acknowledgment

This work was supported by the U.S. National Institutes of Health (NIH), National Cancer Institute (NCI) for the Multiethnic Cohort Study (U01 CA164973 to L.L.M., L.R.W., and C.A.H.); Adiposity Phenotype Study (P01 CA168530 to L.L.M.); and University of Hawaii Cancer Center (P30 CA071789 for Analytical Biochemistry, Biostatistics, Genomics, and Nutrition Support services); and the National Center for Advancing Translational Science, NIH, for recruitment activities at USC (UL1TR000130 to Southern California Clinical and Translational Science Institute). S.A.S. was funded by an NCI training grant (T32 CA229100), a University of Hawaii Cancer Center pilot grant (P30CA071789), and a Multiethnic Cohort Study Supplement (U01 CA164973-13S1). The technical support and advanced computing resources from University of Hawaii Information Technology Services – Research Cyberinfrastructure, funded in part by the National Science Foundation CC* awards # 220148 and #2232862, are gratefully acknowledged. The funders had no role in study design, data collection and analysis, decision to publish, or preparation of the manuscript.

